# The effect of D-cycloserine on brain connectivity over a course of pulmonary rehabilitation

**DOI:** 10.1101/2023.11.05.23298110

**Authors:** Sarah L. Finnegan, Olivia K. Harrison, Martyn Ezra, Catherine J. Harmer, Thomas E. Nichols, Najib M. Rahman, Andrea Reinecke, Kyle T.S. Pattinson

**Affiliations:** Wellcome Centre for Integrative Neuroimaging and Nuffield Division of Anaesthetics, Nuffield Department of Clinical Neurosciences, University of Oxford, Oxford, UK; School of Psychology, University of Otago, Dunedin, New Zealand; Department of Psychiatry, Medical Sciences, University of Oxford, Oxford, UK; Oxford Health NHS Foundation Trust, Warneford Hospital Oxford; Oxford Big Data Institute, Li Ka Shing Centre for Health Information and Discovery, Nuffield Department of Population Health, University of Oxford, Oxford OX3 7LF UK; Nuffield Department of Medicine, University of Oxford, Oxford, UK; Oxford NIHR Biomedical Research Centre, Oxford OX3 7JX

## Abstract

**Rationale:** Combining traditional therapies such as pulmonary rehabilitation with brain-targeted drugs may offer new therapeutic opportunities for the treatment of chronic breathlessness. Recent work has shown that D-cycloserine, a partial NMDA-receptor agonist which has been shown to enhance cognitive behavioural therapies, modifies the relationship between breathlessness related brain activity and breathlessness anxiety over pulmonary rehabilitation. However, whether these changes are supported by alterations to underlying brain structure remains unknown. Here we examine the effect of D-cycloserine over a course of pulmonary rehabilitation on regional brain volume and connectivity.

**Methods:** 72 participants with mild-to-moderate COPD took part in a longitudinal study in parallel to their pulmonary rehabilitation course. Diffusion tensor brain imaging, self-report questionnaires and clinical measures of respiratory function were collected at three time points (before, during and after pulmonary rehabilitation). Participants were assigned to 250mg of D-cycloserine or placebo, which they were administered with on four occasions in a randomised, double-blind procedure.

**Results:** Following four sessions of pulmonary rehabilitation, improvements in breathlessness anxiety were linked with increased insula-hippocampal structural connectivity in the D-cycloserine group. No group differences were found following the completion of pulmonary rehabilitation, or in the integrity of structural connectivity.

**Conclusions:** The action of D-cycloserine on brain connectivity appears to be restricted to within a short time-window of its administration. This temporary boost of the brain connectivity of two key regions associated with the evaluation of unpleasantness may support the re-evaluation of breathlessness cues, illustrated improvements in breathlessness anxiety. This work highlights the relevance of targeting breathlessness expectation in pulmonary rehabilitation.

## Introduction

Brain-targeted drugs may offer novel therapeutic opportunities for the treatment of chronic breathlessness. This new field of research builds upon evidence suggesting that active elements of pulmonary rehabilitation, which remains currently the most effective treatment for chronic breathlessness lie, not in improvements to lung function, but in the reappraisal of the sensory experience [1–4]. Reappraisal of breathlessness anxiety and intensity has been linked using neuroimaging techniques to changes in activity across a network of attentional and emotional regulation regions, including anterior cingulate cortex and insula cortex [3].

Recent research has examined whether the brain associated mechanisms of pulmonary rehabilitation, including processes of reappraisal, could be boosted with the drug D-cycloserine [5]. D-cycloserine is a partial N-Methyl-D-Aspartate (NMDA) receptor agonist, which, acting via glutamateric receptors is thought to increase synaptic plasticity and boost the learning processes associated with exposure-based cognitive behavioural therapies (CBT), such as pulmonary rehabilitation. Considering that 30% of patients undergoing pulmonary rehabilitation derive no clinical benefit [6, 7], and health-related benefits typically return to pre-rehabilitation levels 12-18 months after course completion [8, 9], therapeutic adjuncts such as D-cycloserine are of considerable clinical relevance.

Though brain activity patterns offer a window into short term processes and associated network dynamics in specific situations, they provide less information regarding potential long-term or more general brain changes. Thus, it remains unclear as to whether changes to functional brain activity associated with D-cycloserine administration and subsequent reappraisal of breathlessness anxiety over pulmonary rehabilitation are related to any longer-term structural changes across the brain.

While task based functional brain activity has been shown to be somewhat variable across sessions and individuals [10], structural brain changes, which can be measured in terms of volume or connectivity, have good test-retest reliability and may be more sensitive to experience-dependent long-term changes [11]. Such changes may relate to clinical outcomes.

Measures of regional brain volume and connectivity have been shown to change following exposure-based CBT [12, 13], while posterior and anterior cingulate cortex volume reduction has been linked to symptom improvement (in post-traumatic stress disorder (PTSD)) and successful cognitive reappraisal following CBT [14, 15]. Such structural changes may become therapeutic focuses for brain-targeted drugs. For example, the NMDA receptor blocker, ketamine, has been shown to block corticosteroid induced volume loss within the hippocampus [16]. Potential targets are not limited to NMDA-related pathways; Lithium has also been shown not only to provide symptomatic relief, but also to increase global grey matter volume in patients with bipolar mood disorder [17].

Given that learning is associated with structural changes within grey [18] and white matter [19] and D-cycloserine is thought to boost the learning related process of exposure-based CBT via increased synaptic plasticity, we hypothesised that D-cycloserine would modify brain volume and/or connectivity within/between five key brain regions of interest. These regions were: anterior insular cortex, posterior insular cortex, anterior cingulate cortex, amygdala and hippocampus. Both amygdala and hippocampus have been identified by animal studies as key target sites for the modulation of emotional learning by D-cycloserine [20, 21], while changes to activity within insula and anterior cingulate cortex have been shown to correlate with improvements across pulmonary rehabilitation [3, 5].

## Methods

An overview of the methodology is presented here. Full details can be found within supplementary materials. The study and statistical analysis plan were pre-registered on clinicaltrials.gov (ID: NCT01985750) prior to unblinding. This was an analysis of data from a longitudinal experimental medicines study of patients with COPD over a course of pulmonary rehabilitation. Parts of this study were first published in a characterisation of baseline patient clusters [22] and subsequently in an investigation of the effect of D-cycloserine on functional brain activity across pulmonary rehabilitation [5]. The analysis conducted here is novel and not previously reported.

72 participants (18 female, median age 71 years (46-85 years)) (Table 1 and Figure 1) with COPD were recruited immediately prior to their enrolment in a National Health Service- prescribed course of pulmonary rehabilitation. Written informed consent was obtained from all participants prior to the start of the study. Study approval was granted by South Central Oxford REC B (Ref: 118784, Ethics number: 12/SC/0713). Study inclusion criteria were: a diagnosis of COPD and admittance to pulmonary rehabilitation. Exclusion criteria were: inadequate understanding of verbal and written English, significant cardiac, psychiatric (including depression under tertiary care) or metabolic disease (including insulin-controlled diabetes), stroke, contraindications to either D-cycloserine (including alcoholism) or magnetic resonance imaging (MRI), epilepsy, claustrophobia, regular therapy with opioid analgesics or home oxygen therapy.

**Figure 1.**
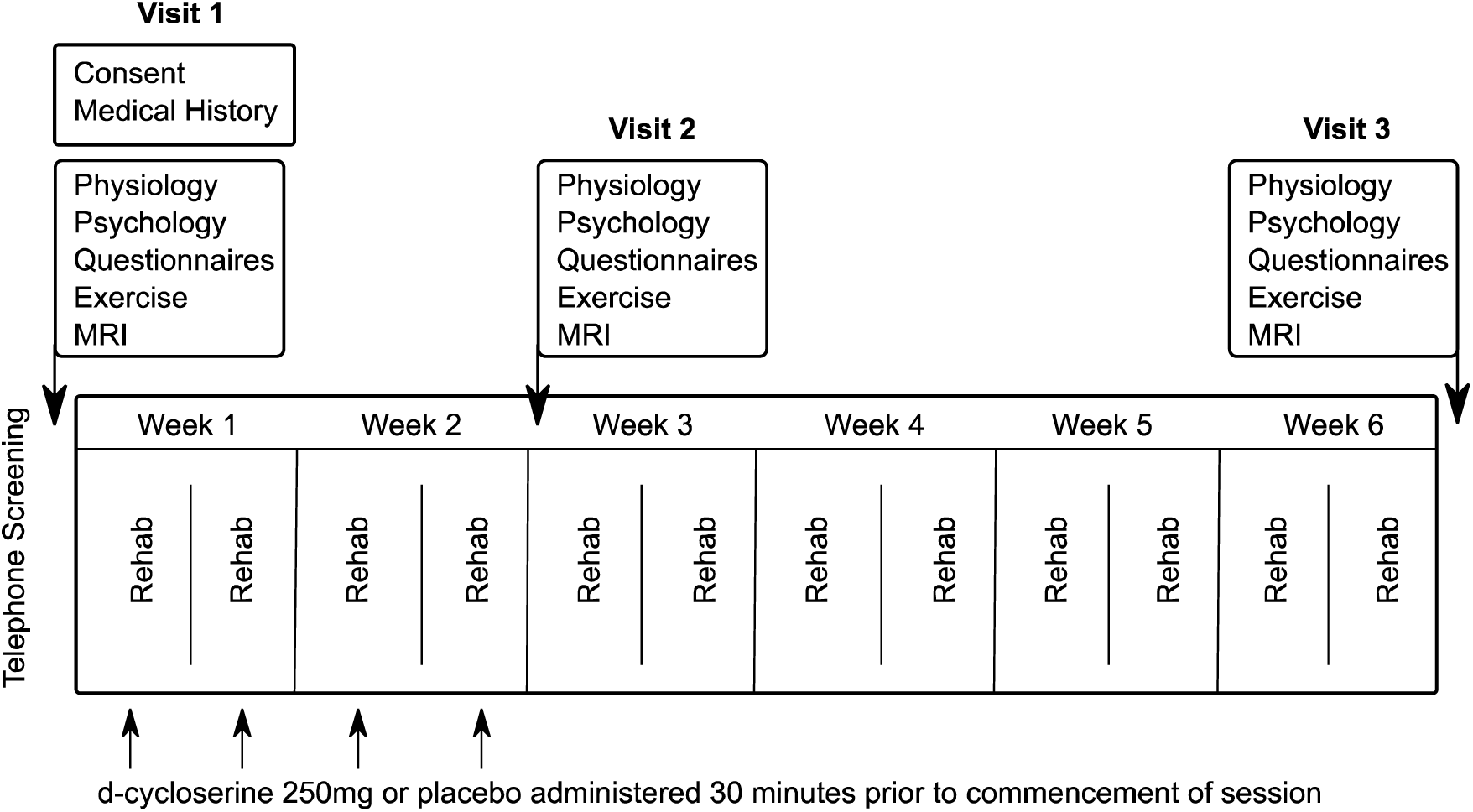
A schematic demonstrating order of visits, rehabilitation sessions and tablet administration throughout the study period. Participants took part in one study visit prior to their first pulmonary rehabilitation session. Study drug/placebo were administered on four occasions over the first four rehabilitation sessions. A second study visit occurred after the final drug/placebo administration. Participants continued with their pulmonary rehabilitation course for a further four weeks before returning for a third study visit.

**Figure 2.**
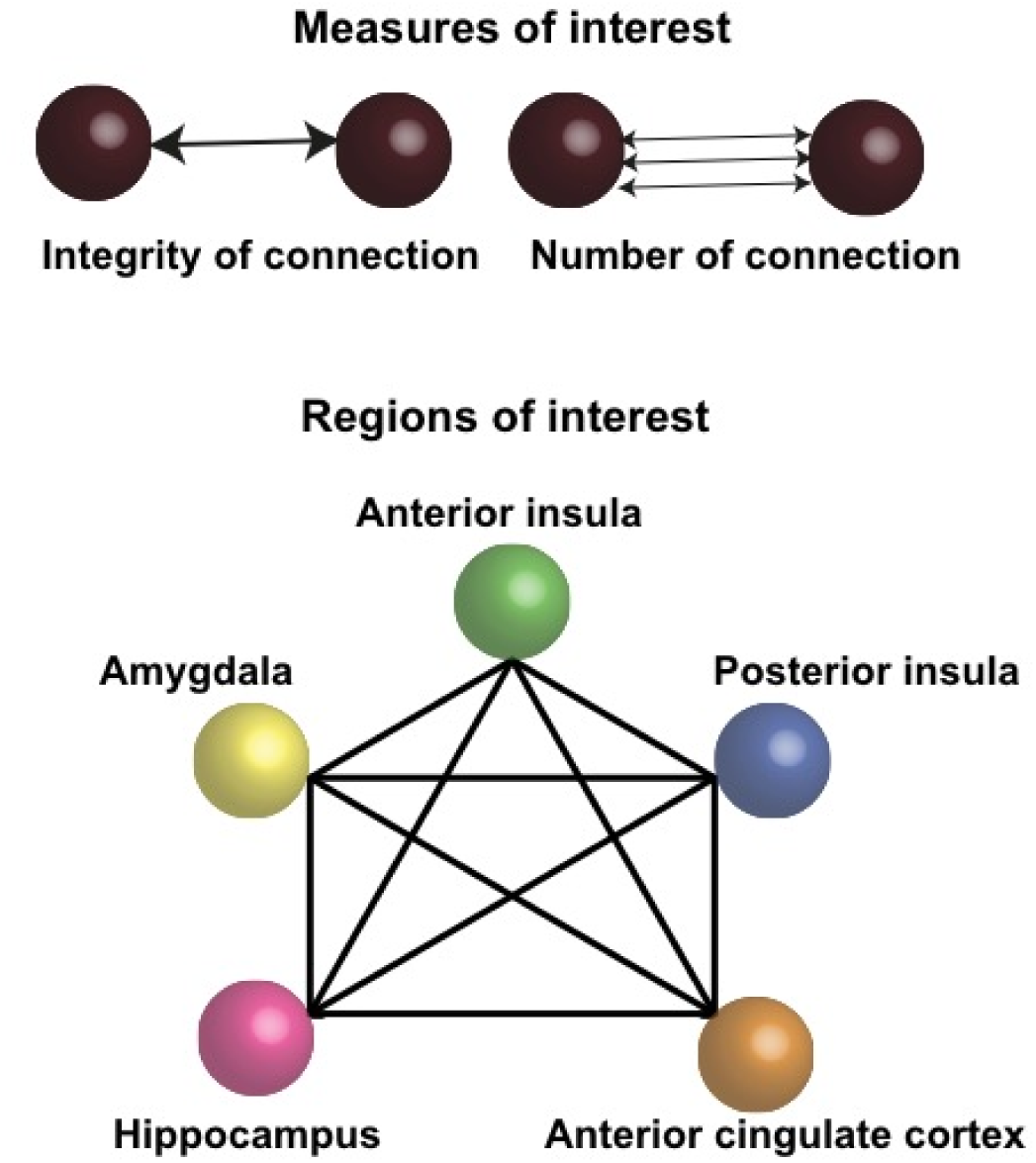
The two measures of interest – the integrity of the regional connections (FA) and the number of connections (streamlines). Schematic representation of the five key regions of interest and their connectivity.

**Table 1.**
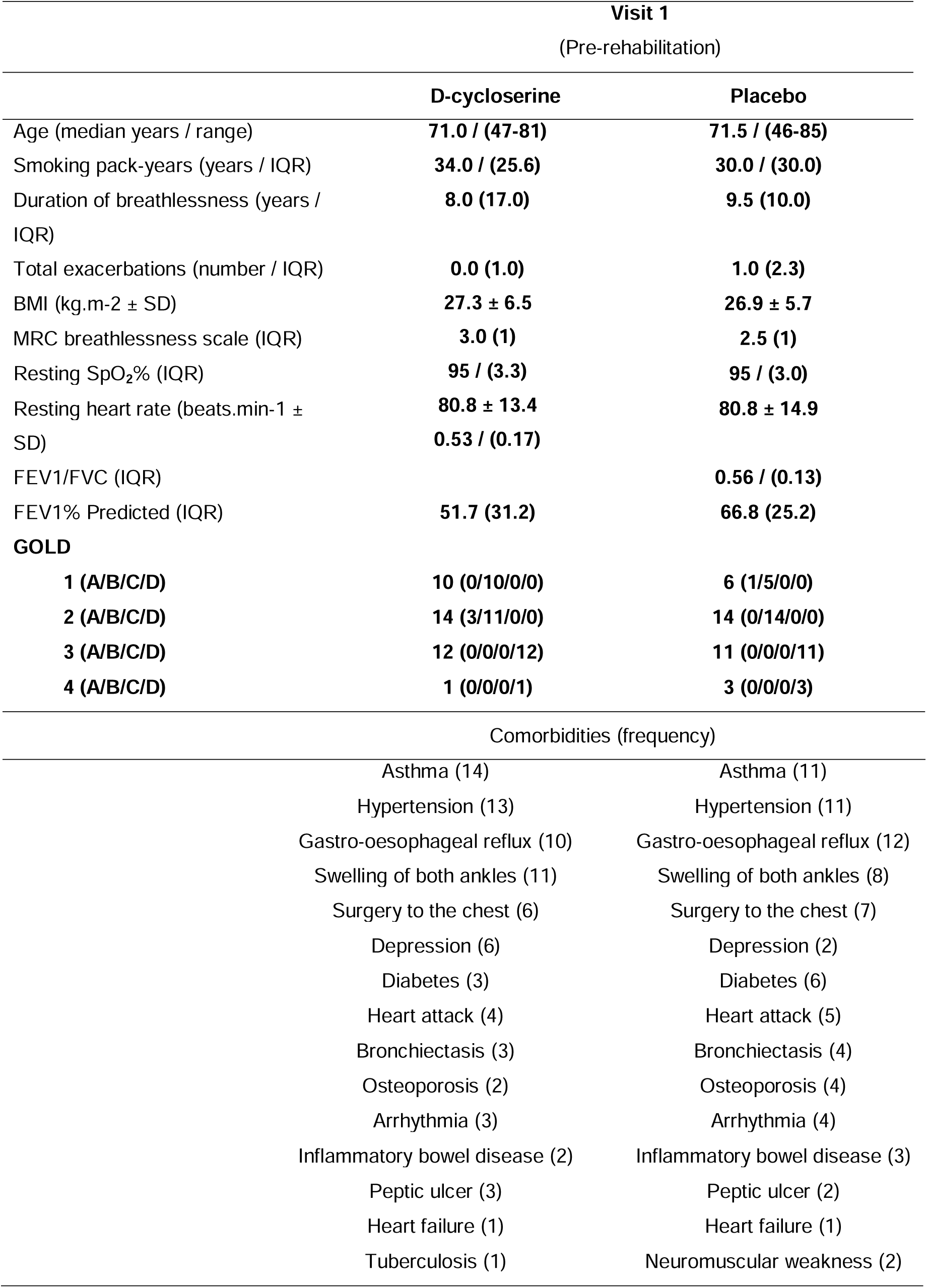
Demographic information from the 72 participants who completed all three study visits. Variance is expression either in terms of standard deviation (SD) or interquartile range (IQR) depending on the normality of the underlying data distribution. **BMI** = Body Mass Index, **MRC** = Medical Research Council. **SpO_2_%** = Peripheral Oxygen saturation, expressed as a percentage. **FEV** = Forced Expiratory Volume. **FVC** = Forced Vital Capacity. Also listed with prevalence in brackets are recorded comorbidities ordered by frequency

### Study drug

Participants were randomised in a double-blinded procedure to receive either 250mg oral D-cycloserine or a matched placebo, administered by the study nurse 30 minutes prior to the onset of their first four pulmonary rehabilitation sessions. Study participants, investigators and those performing the analysis were blinded to the treatment allocation. Both D-cycloserine and placebo were over-encapsulated to appear identical. Full study drug description, randomisation protocol and minimisation criteria can be found within the supplementary materials.

### Study visit protocol

Following telephone screening, participants were invited to attend their first research session (baseline) prior to starting pulmonary rehabilitation. A second study visit took place following the fourth pulmonary rehabilitation session but before the sixth session. Participants completed the remainder of their pulmonary rehabilitation course before attending a third study session (Figure 1) that occurred as close to the termination of pulmonary rehabilitation as possible and always within two weeks.

### Pulmonary rehabilitation

Pulmonary rehabilitation courses were run by either Oxford Health NHS Foundation Trust, West Berkshire NHS Foundation Trust, or Milton Keynes University Hospitals NHS Trust. The full course ran for 6 weeks with two sessions per week including an hour of exercises and an hour of education, as part of a standard pulmonary rehabilitation programme.

### Physiological measures

Spirometry and two Modified Shuttle Walk Tests (MSWT) were collected using standard protocols [23, 24]. Participant height and weight were recorded at each visit. Oxygen saturations and heart rate were measured with pulse oximetry and were collected at rest and following the MSWT.

### MRI Measures

*Image acquisition:* Magnetic resonance imaging of the brain was carried out using a Siemens 3T MAGNETOM Trio. A T1-weighted (MPRAGE) structural scan (voxel size: 1 x 1 x 1 mm) was collected and used for registration purposes. Diffusion weighted images were acquired in the transverse plane using an echo planar imaging sequence (2 acquisitions of 64 directions with 4 non-diffusion weighted images, *b-*value 1,500 s mm^-2^ voxel size 2 x 2 x 2 mm, 64 slices)

A T2*-weighted, gradient echo planar image (EPI) scan sequence (voxel size: 3 x 3 x 3 mm), TR, 3000ms; TE 30ms was used to collect FMRI data. FMRI data are reported elsewhere [5, 25].

### Pre-processing

*Diffusion MRI:* Data were pre-processed following standard steps within FMRIB’s diffusion toolbox (FDT) [26]. First non-brain tissue was removed, and susceptibility induced distortions and eddy currents were corrected using the FSL tools topup and eddy [27, 28]. A diffusion tensor was fitted for each voxel using the FSL tool dtifit. A ball and stick probabilistic model was fitted to the data to estimate tract paths using the FSL tool BEDPOSTX. To estimate the integrity of connections (termed fractional anisotropy (FA)) we followed FSL’s standard Tract Based Spatial Statistics (TBSS) pipeline [27, 29].

#### Connectivity analysis

Probabilistic tractography was run on the output of the probabilistic model with seeds specified as each of the regions of interest (Using the FSL tool PROBTRACKX). A 5x5 matrix of the 5 regions of interest was included, indicating the seed to target masks. This generated values of the total number of streamlines between each region of interest for each subject.

#### Structural integrity analysis - Fractional Anisotropy

All subjects’ FA data were aligned into a common space using the nonlinear registration tool FNIRT [30], which uses a b-spline representation of the registration warp field [31]. Next, the mean FA image was created and thinned to create a mean FA skeleton which represents the centres of all tracts common to the group. Each subject’s aligned FA data was then projected onto this skeleton and the resulting data fed into voxelwise cross-subject statistics. This was carried out using FSL’s TBSS pipeline. To calculate FA values within our five regions of interest an average streamline density map was created by adding together all subjects streamline density maps, created by FSL’s FDT toolbox as part of the streamline analysis. Threshold was set manually to the tightest value for which all regions still connected.

### Group Level Analysis

#### Primary analysis – Focused region of interest analysis

For each patient, the following metrics were extracted from each of the five regions of interest at visits two and three (Supplementary figure 1, panel A).

1. Region to region connectivity (number of streamlines) calculated using probabilistic tractography
2. Regional Fractional Anisotropy (FA), a measure of structural integrity

To test for an overall drug effect across each metric, values relating to regional connectivity and integrity were entered into independent linear mixed effects models where they were adjusted for age and gender. Additionally, connectivity or integrity (FA) at visit one (depending on the metric) was also accounted for within the model. To correct for multiple comparisons across regions, permutation testing (with Family Wise Error Rate (FWE) 5%) was carried out. This process was repeated separately for data collected at visit three. Permutation testing was performed with threshold free cluster enhancement (TFCE) (a non-parametric test) [32] using FSL’s Randomise tool [33] at family wise error corrected p<0.05. The process was then repeated separately for data collected at visit three. To test for whether drug effect was related to improvements in reported breathlessness anxiety, models were run including breathlessness anxiety as an additional model term. Models were programmed using the lme4 function within R version 3.6.1 (2019-07-05).

## Results

There was no significant overall effect of D-cycloserine on mean brain volume (family wise error rate values p>0.05 for the five regions) or connectivity within the five key regions of interest (anterior cingulate, anterior insula cortex, amygdala, hippocampus or posterior insula cortex) (Table 2).

**Table 2.** Significance of overall effect of D-cycloserine on the integrity, measured as fractional anisotropy, and number of connections between the five key regions of interest at visits two and three, having accounted for these measures at visit one. Significance is reported as Family Wise Error (p<0.05) corrected p-values of the difference.

| *Region of interest* | *Visit two (p-value)* |  | *Visit three (p-value)* |  |
| --- | --- | --- | --- | --- |
|  | *Integrity of connections* | *Number of connections* | *Integrity of connections* | *Number of connections* |
| *Anterior cingulate – Anterior insula* | 1.00 | 0.84 | 1.00 | 1.00 |
| *Anterior cingulate – Amygdala* | 0.98 | 0.59 | 0.69 | 0.98 |
| *Anterior cingulate – Hippocampus* | 1.00 | 0.62 | 0.34 | 1.00 |
| *Anterior cingulate – Posterior insula* | 1.00 | 0.89 | 0.97 | 1.00 |
| *Anterior insula - Amygdala* | 0.99 | 1.00 | 0.98 | 0.94 |
| *Amygdala – Posterior insula* | 1.00 | 1.00 | 0.97 | 0.85 |
| *Anterior insula - Hippocampus* | 1.00 | 0.54 | 0.98 | 1.00 |
| *Amygdala – Hippocampus* | 1.00 | 1.00 | 0.97 | 0.91 |
| *Anterior insula – Posterior insula* | 1.00 | 1.00 | 0.69 | 0.77 |
| *Hippocampus – Posterior insula* | 0.24 | 0.85 | 0.98 | 0.95 |

A positive covariance was observed between the number of connections of anterior insula and hippocampus and change in breathlessness anxiety in the D-cycloserine group at visit two (p=0.04, corrected family wise error rate 5%) compared to the placebo group (Table 2). This relationship was such that participants who gave lower ratings for breathless anxiety had a correspondingly higher number of connections between the two regions. This effect was no longer observed at visit three, following the completion of pulmonary rehabilitation. No differences were observed in fractional anisotropy or volume of any region of interest at visit two or three.

No differences were found in changes to reported breathlessness anxiety between D- cycloserine and placebo groups at visits two or three (Table 3).

**Table 3.** Significance of the relationship between D-cycloserine and breathlessness anxiety (wA) on the integrity, measured as fractional anisotropy and number of connections between the five key regions of interest at visits two and three, having accounted for these measures at visit one. Significance is reported as Family Wise Error (p<0.05) corrected p-values of the difference.

| Region of interest | Visit two (p-value) allowing<br>for an interaction with wA |  | Visit three (p-value) allowing<br>for an interaction with wA |  |
| --- | --- | --- | --- | --- |
|  | Integrity of<br>connections | Number of<br>connections | Integrity of<br>connections | Number of<br>connections |
| Anterior cingulate – Anterior insula | 0.99 | 0.96 | 1.00 | 1.00 |
| Anterior cingulate – Amygdala | 0.89 | 0.99 | 0.98 | 1.00 |
| Anterior cingulate – Hippocampus | 0.99 | 0.12 | 0.84 | 1.00 |
| Anterior cingulate – Posterior insula | 0.90 | 0.99 | 0.99 | 0.98 |
| Anterior insula - Amygdala | 0.50 | 0.99 | 0.96 | 1.00 |
| Amygdala – Posterior insula | 0.75 | 0.99 | 1.00 | 1.00 |
| **Anterior insula - Hippocampus** | 0.99 | **0.04\*** | 0.98 | 1.00 |
| Amygdala – Hippocampus | 0.99 | 0.99 | 0.98 | 1.00 |
| Anterior insula – Posterior insula | 0.99 | 0.99 | 0.99 | 1.00 |
| Hippocampus – Posterior insula | 0.83 | 0.99 | 0.99 | 0.95 |

**Table 4.** Scores on breathlessness anxiety for drug and placebo group during (visit two) and following (visit three) pulmonary rehabilitation. Measures are expressed as a “change score”. Values are calculated as baseline visit (visit one) minus the later visit, thus positive values indicate an improvement. Significance is reported as Family Wise Error (p<0.05) corrected p-values of the difference in scores between drug and placebo groups at visit two, accounting for scores at visit one. Values are either recorded as mean and standard deviation (1) or as median and interquartile range (2) depending on the normality of distribution

|  | D-cycloserine | Placebo | p-value |
| --- | --- | --- | --- |
| Visit 2 |  |  |  |
| Breathlessness Anxiety (wA) | **0.4 (29.0)<sup>2</sup>** | **5.0 (17.8)<sup>2</sup>** | **0.15** |
| Visit 3 |  |  |  |
| Breathlessness Anxiety (wA) | 3.0 (7.0)<sup>2</sup> | 7.3 (16.8)<sup>2</sup> | 0.053 |

## Discussion

### Key Findings

Our results showed that by visit two (after four sessions of rehabilitation), participants who received D-cycloserine and reported lower breathlessness anxiety had a greater connectivity between anterior insula and hippocampus. Given that both regions are associated with the evaluation of unpleasantness [34], we suggest that this change in connectivity may relate to plasticity associated with a re-evaluation of the situations described by the breathlessness- cues. By visit three (after the completion of pulmonary rehabilitation), this effect was no longer observed, suggesting that this relationship may be temporally linked to the D-cycloserine administration or to its combination with the early stages of pulmonary rehabilitation. These findings suggest that brain-targeted drugs may have the potential to “temporally-boost” brain changes associated with pulmonary rehabilitation.

Pulmonary rehabilitation draws on elements of exposure-based cognitive behavioural therapy to provide symptomatic relief for around 70% of participants [3, 4, 35]. However, health-related benefits return to pre-rehabilitation levels 12-18 months after course completion [8, 9] for the majority of participants. This has led to speculation as to whether brain-targeted drugs have potential to either boost or extend the overall therapeutic efficacy of pulmonary rehabilitation.

Building on our previous work in which we showed brain activity changes within key attentional regulation brain regions to be associated with improvements in breathlessness anxiety over pulmonary rehabilitation [3, 5], it was hypothesised that shifts in expectation-related brain activity relating to learned breathlessness associations may be augmented by D-cycloserine. The action of D-cycloserine is thought to occur by changing activity within hippocampus, amygdala, dorsolateral prefrontal cortex and insula, regions which overlap with brain networks of internal bodily sensation (interoception) and reappraisal [36–38]. Using a double-blinded placebo-controlled design we were able to test this hypothesis. In the D-cycloserine group, for a given change in ratings of breathlessness anxiety, there was correspondingly less brain activity in response to breathlessness cues than the placebo group across a network of emotional salience regions [5], including superior frontal gyrus, precuneus, dorsolateral prefrontal cortex and medial prefrontal cortex. We concluded that this represented a down-regulation of emotional responses to breathlessness word cues as a result of D-cycloserine in people who had derived positive change from pulmonary rehabilitation. However, questions remained outstanding as to whether brain activity differences were supported by any underlying structural changes.

It is well established that learning processes are associated with changes to brain structure [18, 39], while brain targeted drugs have also been shown to affect both structural and functional connectivity [16, 40], as well as regional volume [17]. Here we identified no overall effect of D-cycloserine either on brain volume or connectivity. Instead, in keeping with our previous findings we found structural changes were related to changes in behavioural anxiety, a surrogate marker of rehabilitation success. This result fits with a growing literature base regarding D-cycloserine, which suggests that D-cycloserine not only makes “good exposures” better but also may make “bad exposures” worse” [41].

Thus, by taking into account improvements to breathlessness anxiety we identified a greater number of connections between anterior insula and hippocampus in the D-cycloserine group for people who reported a greater reduction in breathlessness anxiety after the first four sessions of pulmonary rehabilitation. By the end of pulmonary rehabilitation (visit three) the difference between placebo and D-cycloserine groups was no longer observed, suggesting this effect to be related temporally to the D-cycloserine administration period or its interaction with the initiation of pulmonary rehabilitation. It is possible that the short-term relationship between connectivity and change in breathlessness anxiety may have then facilitated the functional changes observed across a network of emotional salience regions at visit three [5].

The differences in timing of the brain networks targeted by D-cycloserine fits with recent meta-analysis of D-cycloserine action [42] and offers potential insight into the nature of breathlessness reappraisal. It has been suggested that the somewhat unpredictable success rate of D-cycloserine in exposure-based therapies could be explained by whether the target for learning relies on high- or low-order conditioned responses. Lower-order responses are automatic, such as the startle reflex, and rely on autonomic fight-flight responses, whereas higher-order responses describe a reaction learned over time following repeated exposures. Lower-order responses may be easier targets for D-cycloserine’s NMDA-receptor action via the limbic system, which include anterior insula – hippocampus connections, compared to higher-order learned responses embedded within frontal networks [42]. Certainly, breathlessness can be considered as both a high and low order conditioned response. While the experience of breathlessness lies within the autonomic, implicit processing, for many the deeply embedded and individual learned associations will be higher-order and more resilient to D-cycloserine action. Thus, as we observe here, the first sites of action for D-cycloserine may be measured at anterior insula and hippocampus, and result in the revaluation of the unpleasantness of breathlessness. This effect then translates over the course of pulmonary rehabilitation to tackle the more deeply embedded fearful associations of breathlessness, resulting in broader functional changes across higher-order brain regions: superior frontal gyrus, precuneus, dorsolateral prefrontal cortex and medial prefrontal cortex.

## Limitations and future directions

Our findings suggest that D-cycloserine action relates to the success of pulmonary rehabilitation. While we were able to use changes to breathlessness anxiety as a surrogate marker for pulmonary rehabilitation success, future studies should consider collecting post-session reports for model inclusion. Questions remain outstanding as to why this effect was seen at visit two (after four sessions of rehabilitation) but not at visit three (after the completion of pulmonary rehabilitation). Given that there was no overall group difference in the number of connections between the two groups between any of the regions or in scores of breathlessness anxiety, we can conclude that it is not as a result of the number of connections or rated behavioural scores at visit three “catching up” with those at visit two. Additionally, pulmonary rehabilitation is a strong behavioural intervention, and recent literature has suggested that D-cycloserine action is curtailed near the therapeutic ceiling [43]. Thus, pairing a strong drug with a weaker behavioural therapy may provide more scope for mechanistic investigations of overall group differences. However, the importance of rehabilitation outcome on changes to brain connectivity highlights the value of accounting for effects of therapy on the individual. With this more sensitive approach we were able to detect drug induced brain changes which would not have been apparent using traditional clinical endpoints. Future studies should consider accounting for individual differences with more formally specified outcome measures.

## Conclusions

We have provided evidence suggesting that D-cycloserine may have a temporary effect on brain structure, producing increased connectivity between anterior insula and hippocampus in people who report greater changes in breathlessness after four sessions of pulmonary rehabilitation. It is possible that these changes may be a precursor for changes to brain activity and the reappraisal of emotional responses to breathlessness related cues. This effect may help us unpick the mechanism of effect for pulmonary rehabilitation and could be extended to other brain-targeted drugs in the future.

## Data Availability

All data produced in the present study are available upon reasonable request to the authors

## Author contributions

**S.L.F –** Acquisition of data, analysis, interpretation, drafting, editing and approving manuscript

**O.K.H - I**nterpretation, editing and approving manuscript

**M.E** – Analysis, editing and approving manuscript

**C.J.H –** Study design, editing and approving manuscript

**T.N** – Analysis, approving manuscript

**N.M.R -** Study design, editing and approving manuscript

**A.R -** Study design, editing and approving manuscript

**K.T.S.P -** Study design, interpretation, editing and approving manuscript

